# Antibiotic Resistance associated with the COVID-19 Pandemic: A Rapid Systematic Review

**DOI:** 10.1101/2022.09.01.22279488

**Authors:** BJ Langford, J-PR Soucy, V Leung, M So, ATH Kwan, JS Portnoff, S Bertagnolio, S Raybardhan, D MacFadden, N Daneman

## Abstract

**Background:** COVID-19 and antimicrobial resistance (AMR) are two intersecting global public health crises.

**Objective:** We aim to describe the impact of the COVID-19 pandemic on AMR across healthcare settings.

**Data Source:** A search was conducted in December 2021 in World Health Organization’s COVID-19 Research Database with forward citation searching up to June 2022.

**Study Eligibility:** Studies evaluating the impact of COVID-19 on AMR in any population were included and influencing factors were extracted.

**Methods:** Pooling was done separately for Gram-negative and Gram-positive organisms. Random effects meta-analysis was performed.

**Results:** Of 6036 studies screened, 28 were included and 23 provided sufficient data for meta-analysis. The majority of studies focused on hospital settings (n=25, 89%). The COVID-19 pandemic was not associated with a change in the incidence density (IRR 0.99, 95% CI: 0.67 to 1.47) or proportion (RR 0.91, 95% CI: 0.55 to 1.49) of MRSA or VRE cases. A non-statistically significant increase was noted for resistant Gram-negatives (i.e., ESBL, CRE, MDR or carbapenem-resistant *Pseudomonas* or *Acinetobacter* species, IRR 1.64, 95% CI: 0.92 to 2.92; RR 1.08, 95% CI: 0.91 to 1.29). The absence of enhanced IPAC and/or ASP initiatives was associated with an increase in Gram-negative AMR (RR 1.11, 95%CI: 1.03 to 1.20), while studies that did report implementation of these initiatives noted no change in Gram-negative AMR (RR 0.80, 95%CI: 0.38 to 1.70). However, a test for subgroup differences showed no statistically significant difference between these groups (P=0.40)

**Conclusion:** The COVID-19 pandemic could play an important role in the emergence and transmission of AMR, particularly for Gram-negative organisms in hospital settings. There is considerable heterogeneity in both the AMR metrics utilized and the rate of resistance reported across studies. These findings reinforce the need for strengthened infection prevention, antimicrobial stewardship, and AMR surveillance in the context of the COVID-19 pandemic.

**PROSPERO registration: CRD42022325831:** This research was carried out as part of routine work, no funding was received Data collection template, data, and analytic code are available upon request.

## Background

High antibiotic use in patients with COVID-19 threatens to contribute to the antimicrobial resistance crisis. Although antibiotics do not treat COVID-19, they are commonly used because of initial diagnostic uncertainty in patients presenting with respiratory illness, and of concern for bacterial co-infection or secondary infection in those with confirmed COVID-19. We performed a series of rapid reviews as part of a living review on concomitant bacterial infection in the context of COVID-19 and found high antibiotic prescribing (approximately 75%) despite the relatively low bacterial infection rates, particularly in patients outside of the ICU setting (<10%).^1–3^

Our most recent systematic review identified COVID-19 patients as a potential important reservoir for antimicrobial resistance. Over 60% of patients with COVID-19 who had a bacterial infection were infected with a highly resistant organism.^4^ Due to person-to-person transmission of organisms, particularly in healthcare settings, this presents a threat to the broader population beyond those with COVID-19.

While substantial inappropriate antibiotic prescribing has occurred in patients with COVID-19, antibiotic use for other infectious syndromes has declined early in the pandemic, particularly in community settings.^5,6^ This could potentially be due to the attenuation of transmission of other viral and bacterial pathogens due to public health measures to contain COVID-19, including physical distancing and masking. Enhanced infection prevention and control activities in healthcare settings could further mitigate the impact on AMR.^7^ Given potentially opposing effects, it is unclear how selection of antibiotic resistance in bacteria has occurred across populations during the pandemic. Emerging data from the United States Centers for Disease Control and Prevention suggests the pandemic has resulted in rising rates of AMR, including carbapenem-resistant *Acinetobacter* and extended spectrum beta-lactamase producing *Enterobacterales*.^8^

While we have reported that antimicrobial resistance is high in individual patients with COVID-19 and bacterial infection, the ecological impact of the pandemic on AMR at the population level is not yet well-described. We aim to describe the impact of the COVID-19 pandemic on antimicrobial resistance across healthcare settings.

## Methodology

### Searches

We performed a comprehensive search of the World Health Organization (WHO) COVID-19 Research Database for published literature in any language from January 1, 2019 to December 1, 2021 with assistance from medical library information specialists. The WHO COVID-19 Research Database is a comprehensive multilingual source of COVID-19 literature updated weekly that includes citations from Medline, Scopus, CINAHL, ProQuest Central, Embase, and Global Index Medicus.^9^ The search strategy was structured to include co-infection or secondary infection terms and bacterial infection terms which were applied to the COVID-19 literature in the database. The full search strategy is available in the supplement. Forward citation searching was performed in Google Scholar to capture more recent publications up to June 2022.^10^

### Study Eligibility

All studies in inpatient and outpatient settings were eligible for inclusion. The following inclusion and exclusion criteria were applied:

#### Inclusion Criteria

1. Study provides data on antibiotic resistance before (before January 2020, or as identified by authors) vs. during the COVID-19 pandemic (January 2020 or later, or as identified by authors) in a specific healthcare setting.
2. Antibiotic resistance is reported as 1) incidence density rate (e.g., rate per 1000 patient days or per patient population), and/or 2) effect measure (e.g., risk, odds, rate ratio) of antimicrobial resistance, and/or 3) prevalence of antimicrobial resistant organisms (e.g., methicillin-resistant *Staphylococcus aureus* (MRSA) out of all Staphylococcus aureus).

Antibiotic resistance includes any of the following pathogens, as defined by study authors: MRSA, vancomycin-resistant *enterococci* (VRE), carbapenem or multi-drug resistant (MDR) *Pseudomonas spp*., carbapenem or MDR *Acinetobacter spp*., extended-spectrum beta-lactamase (ESBL)-producing (or third generation cephalosporin resistant) *Enterobacterales*, carbapenem-resistant *Enterobacterales* (CRE).

#### Exclusion Criteria

1. Reviews, editorials, case studies, case series, letters, pre-print publications, dissertations, poster presentations.
2. Studies including <100 patients.
3. Presumed or suspected bacterial infections (rather than confirmed infections).
4. Studies combining bacterial and non-bacterial co-infection as a single metric.

### Population

Individuals receiving care in any healthcare setting and in any age group.

### Main Outcomes

The main outcome is the incidence of antimicrobial resistance in the population associated with COVID-19, either expressed as an incidence density rate (antibiotic resistant infections per 1000 patient days) or proportion (e.g. proportion of *S. aureus* that were MRSA, proportion of patient admissions with resistant infection).

### Data Screening and Extraction

Records were managed using Covidence bibliographic software. All titles and abstracts were screened by a single author (in our previous review,^4^ there was substantial reviewer agreement, kappa: 0.66). Full text screening was performed by at least a single author (in the previous review, we determined kappa to be substantial at 0.62 to 0.68). A single review author extracted study characteristics and data according to a pre-defined list of study elements, with a second check by another review author. Study characteristics including design, patient population, and antimicrobial resistance metrics were extracted. We also extracted whether the authors indicated infection prevention and control (IPAC) measures were strengthened during the pandemic and/or whether there was an antimicrobial stewardship program (ASP) in place. These variables were extracted in order to stratify changes in AMR based on potential AMR-mitigating factors.

### Risk of Bias Assessment

We used a 10-item validated risk of bias in prevalence studies tool incorporated into data extraction (5).

### Data Analysis

Findings were summarized descriptively. In studies providing complete numerator and denominator data, incidence rate ratios (IRR) were pooled using a GLMM random-effects meta-analysis and risk ratios (RR) were pooled using Mantel-Haenszel random effects meta-analysis with between-study variance estimated using the Paule-Mandel estimator.. Results were presented in forest plots and pooled across Gram-positive and Gram-negative organisms, stratified by the reporting of enhanced IPAC measures and/or ASP. All analyses were carried out using R version 4.1.2 with the packages *metafor* and *meta*.

Heterogeneity was assessed using the I^2^ statistic, with <40% considered low heterogeneity, 30–60% considered moderate heterogeneity, 50–90% considered substantial heterogeneity, and 75–100% considered considerable heterogeneity.^11^

## Results

Of 6036 studies identified via literature search, 28 were eligible for inclusion (18 via full-text screening, 9 via forward citation screening, and 1 expert-identified).^12–39^ The most common countries of origin were the United States (n=4), Italy (n=4), and Brazil (n=3). Patient populations studied included all hospitalized patients (n=17), those hospitalized in intensive care units only (n=5), special populations (e.g., oncology, surgery) (n=3), mixed hospitalized and community-dwelling patients (n=2), and community-dwelling patients only (n=1). Studies evaluated a range of both community- and healthcare-acquired infection. Combined healthcare and community-acquired infection or not setting of acquisition not distinguished was most common (n=15), followed by only healthcare-associated (n=11), and only community-acquired (n=2). The majority of studies derived resistance data from clinical specimens (n=20), six included both clinical and screening specimens or did not specify the type of specimen, followed by two studies using screening specimens only. Most studies had moderate risk of bias (n=18), followed by low (n=5), and high (n=5) risk of bias.

### Measures of Antimicrobial Resistance

Incidence density (e.g., cases of resistant infections per 1000 patient days) was most commonly used to measure a change in AMR associated with COVID-19 (n=11) and proportion of isolates or infections (e.g., percentage of *S. aureus* cases that were MRSA, n=11), followed by incidence (e.g., cases per admission or discharges, n=5), and other (e.g. standardized infection ratio, point prevalence n=2). Study details and AMR metric directionality are provided in Table 1. Of the 28 eligible studies, 23 (82%) provided raw numerator and denominator data to facilitate meta-analysis.

**Table 1.**
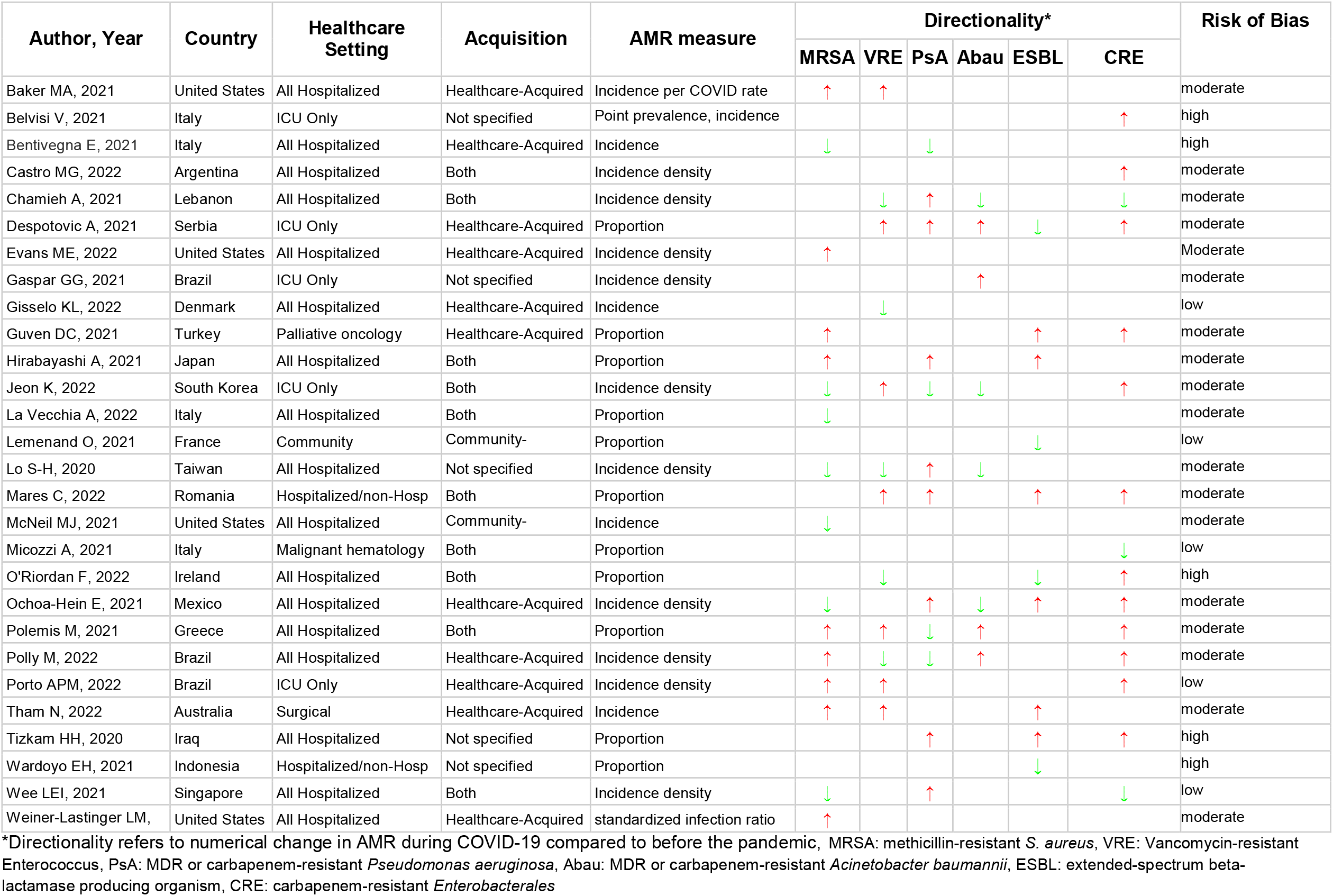
AMR Associated with COVID-19 Study Characteristics.

### Resistance in Gram Positive Organisms

#### MRSA (n=15)

Over 6,848,357 patient days of follow-up, our meta-analysis found that the COVID-19 pandemic was not associated with a change in incidence rate of MRSA (IRR 1.03, 95%CI: 0.65 to 1.62, I^2^=95%, n=5). Similarly the COVID-19 pandemic was not associated with a change in the proportion of cases that were MRSA (RR 0.91, 95%CI: 0.60 to 1.36, I^2^=93%, n=7).

#### VRE (n=12)

Over 356,056 patient days, meta-analysis shows that the COVID-19 pandemic was not associated with a change in the incidence of VRE (IRR 0.75, 95%CI: 0.49 to 1.15, I^2^=56%, n=3). Similarly there was no change in the proportion of VRE cases (RR 0.91, 95%CI: 0.30 to 2.79, I^2^=94%, n=5).

### Overall Gram-Positive Resistance and Association with Infection Prevention and Antimicrobial Stewardship Initiatives

When pooling both MRSA and VRE, no association was found between COVID-19 pandemic and the incidence (IRR 0.99, 95%CI: 0.67 to 1.47, I^2^=91%, n=8) or proportion (RR 0.91, 95%CI: 0.55 to 1.49, I^2^=92%, n=12) of resistant Gram-positive cases. The presence of IPAC or ASP initiatives was not associated with a statistically significant difference resistance rates (with IPAC/ASP: RR 0.59, 95%CI 0.15 to 2.42, I^2^=89%, n=4; without IPAC/ASP: RR: 1.15, 95%CI: 0.94 to 1.41, I^2^=89% n=8, test of subgroup difference P=0.36). See Figure 2 and 3.

**Figure 1.**
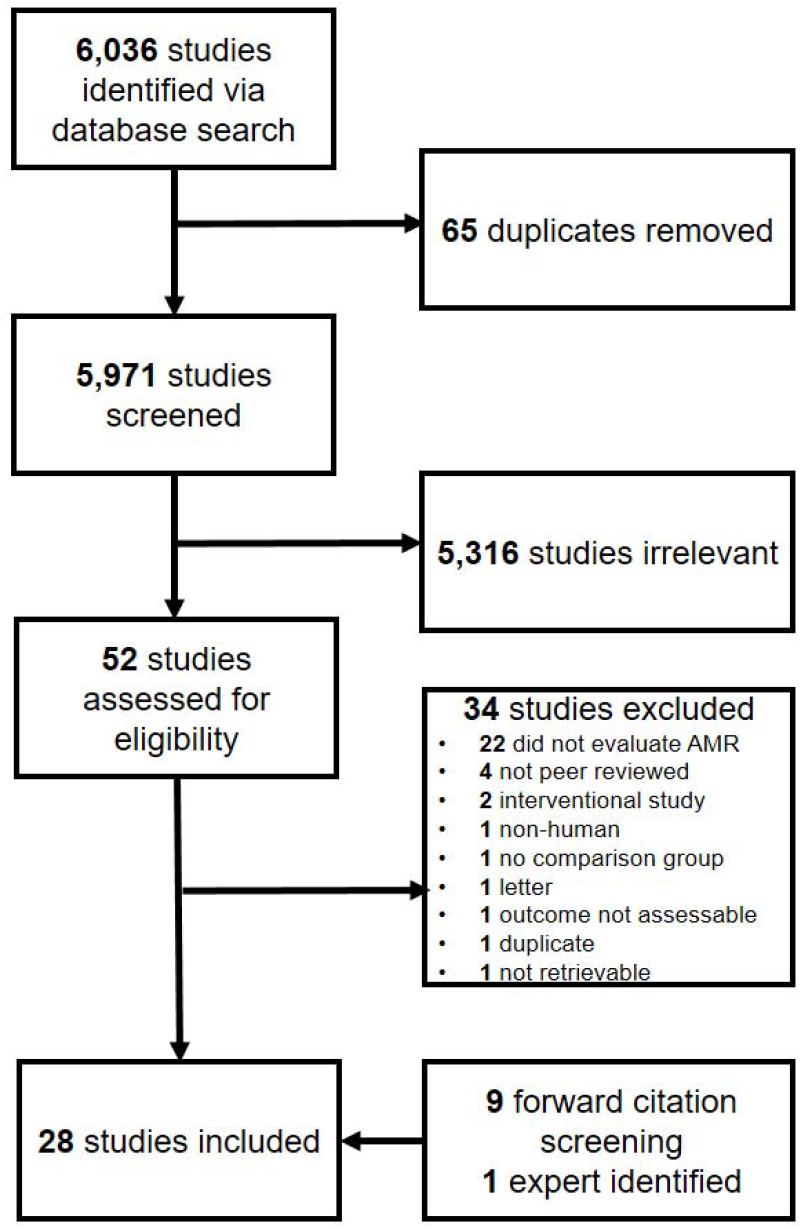
PRISMA Flow Diagram

**Figure 2.**
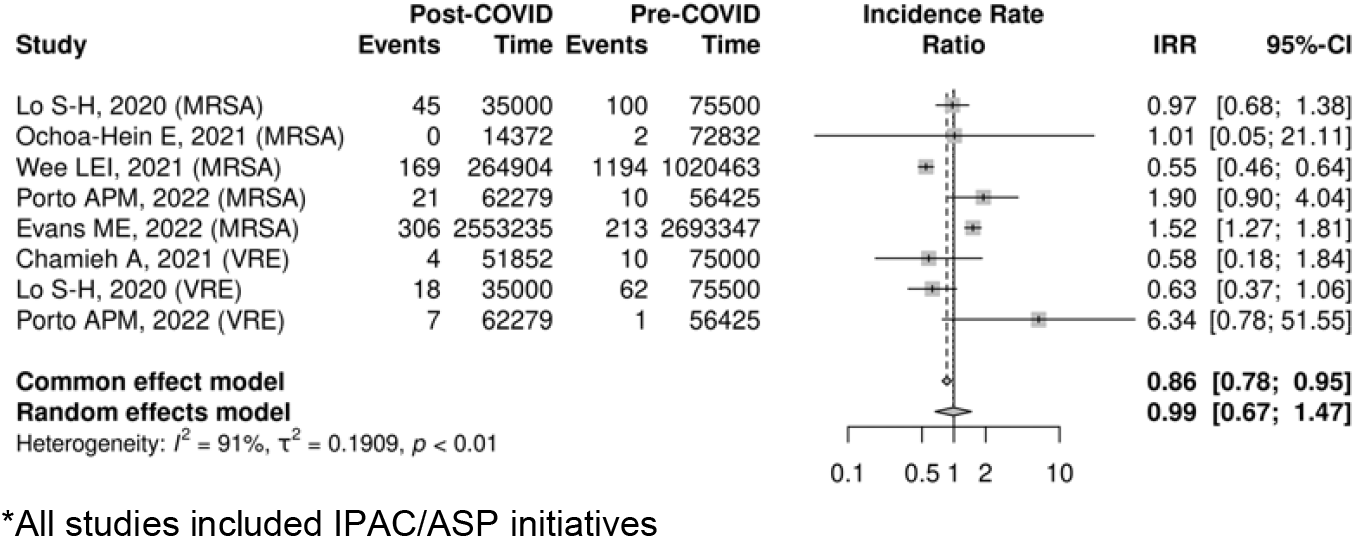
COVID-19 Pandemic and Gram-Positive AMR Incidence Rate Ratio.

**Figure 3.**
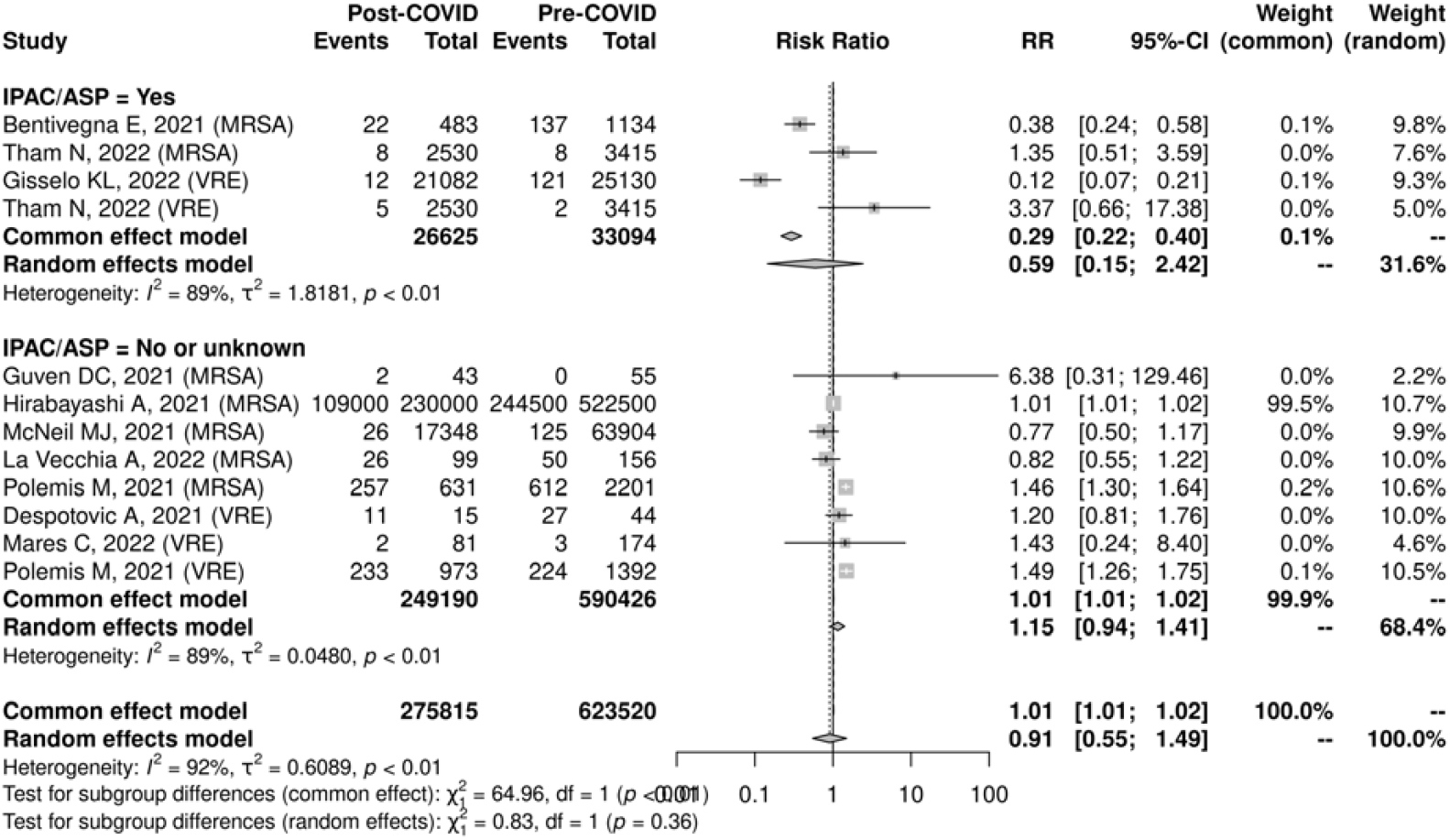
COVID-19 Pandemic and Gram-Positive AMR Risk Ratio.

### Resistance in Gram-Negative Organisms

#### Resistant Acinetobacter spp. (n=8)

Across 325,847 patient days, there was no association between COVID-19 and the incidence of carbapenem- or multi-drug resistant *Acinetobacter* spp. (IRR 0.79, 95%CI: 0.30 to 2.07, I^2^=77%, n=4). However, there was a small increase in the proportion of infections that were resistant *Acinetobacter spp*. (RR 1.02, 95%CI 1.01 to 1.03, I^2^=0%, n=2).

#### Resistant Pseudomonas spp. (n=10)

Across 1,609,923 patient days, there was no association between COVID-19 and the incidence of resistant *Pseudomonas* infections (IRR 1.10, 95%CI 0.91 to 1.30, I^2^=0%, n=4). Similarly there was no association with the proportion of cases that were resistant (RR 1.02, 95%CI: 0.85 to 1.23, I^2^=58%, n=6).

### ESBL (n=10)

One study with 87,204 patient days of follow-up found an increased IRR associated with the COVID-19 pandemic (IRR 15.20, 95%CI: 4.90 to 47.14). However the proportion of cases with an ESBL-producing organism was not significantly altered with COVID-19 (RR: 1.10, 95%CI: 0.91 to 1.33, I^2^=94%, n=8).

### CRE (n=15)

Across 587,047 patient days, there was no significant change detected in the incidence of carbapenem-resistant *Enterobacterales* (*E. coli* and *Klebsiella spp*.) (IRR 2.05, 95%CI: 0.77 to 5.44, I^2^=95%, n=5). Similarly, there was no identified increase in proportion of cases that were CRE (RR 1.10, 95%CI: 0.61 to 1.99, I^2^=88%, n=6).

### Overall Gram-Negative Resistance and Association with Infection Prevention and Antimicrobial Stewardship

When pooling all resistant Gram-negative organisms, there was a non-statistically significant association between COVID-19 pandemic and the incidence rate (IRR 1.64, 95%CI: 0.92 to 2.92, I^2^=93%, n=14) as well as the proportion of cases that were resistant (RR 1.08, 95%CI: 0.91 to 1.29, I^2^=92%, n=22). The absence of reported enhanced IPAC and/or ASP was associated with an increase in Gram-negative AMR (RR 1.11, 95%CI: 1.03 to 1.20, I^2^=88%, n=5), whereas no association was seen in studies that did report such initiatives (RR 0.80, 95%CI: 0.38 to 1.70, I^2^=90%, n=17). However, a test for subgroup differences showed no statistically significant difference between these groups (P=0.40). Figure 4 and 5.

**Figure 4.**
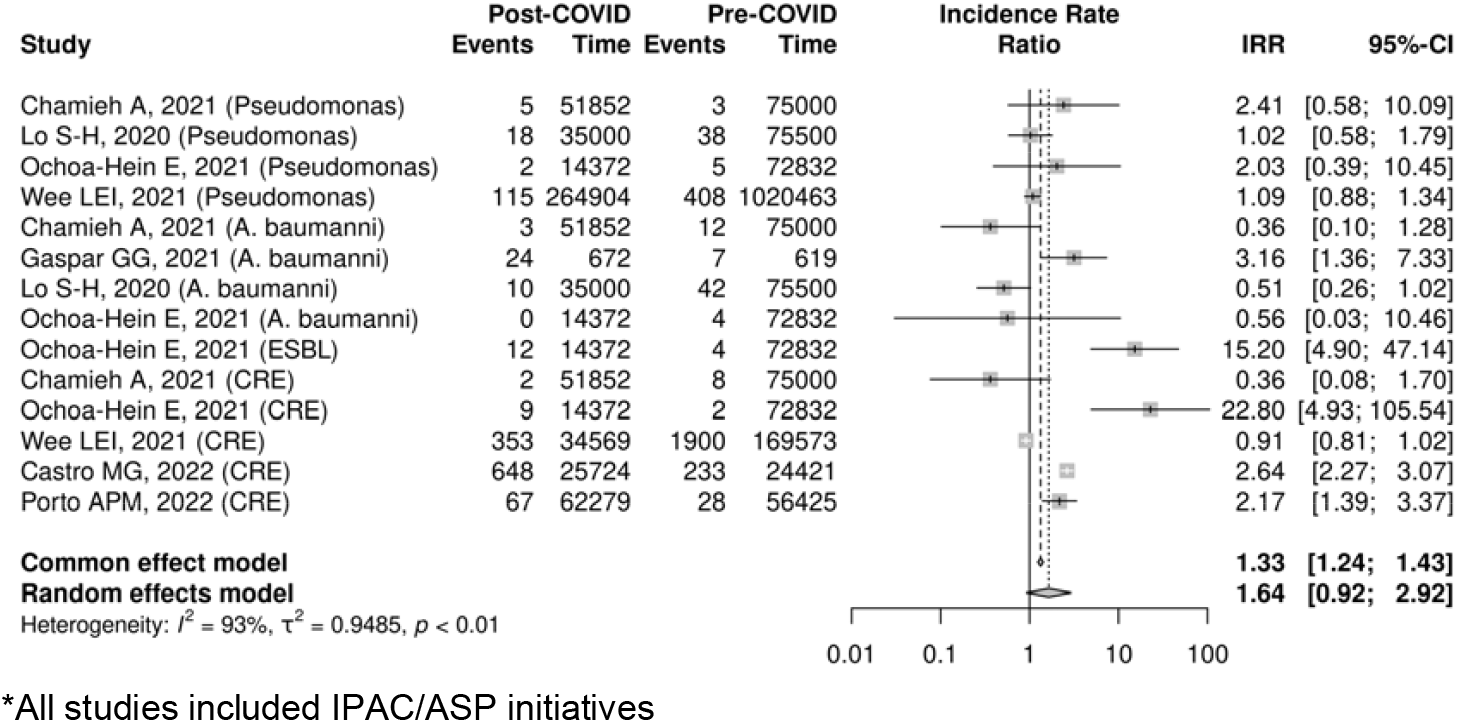
COVID-19 Pandemic and Gram-Negative AMR Incidence Rate Ratio.

**Figure 5.**
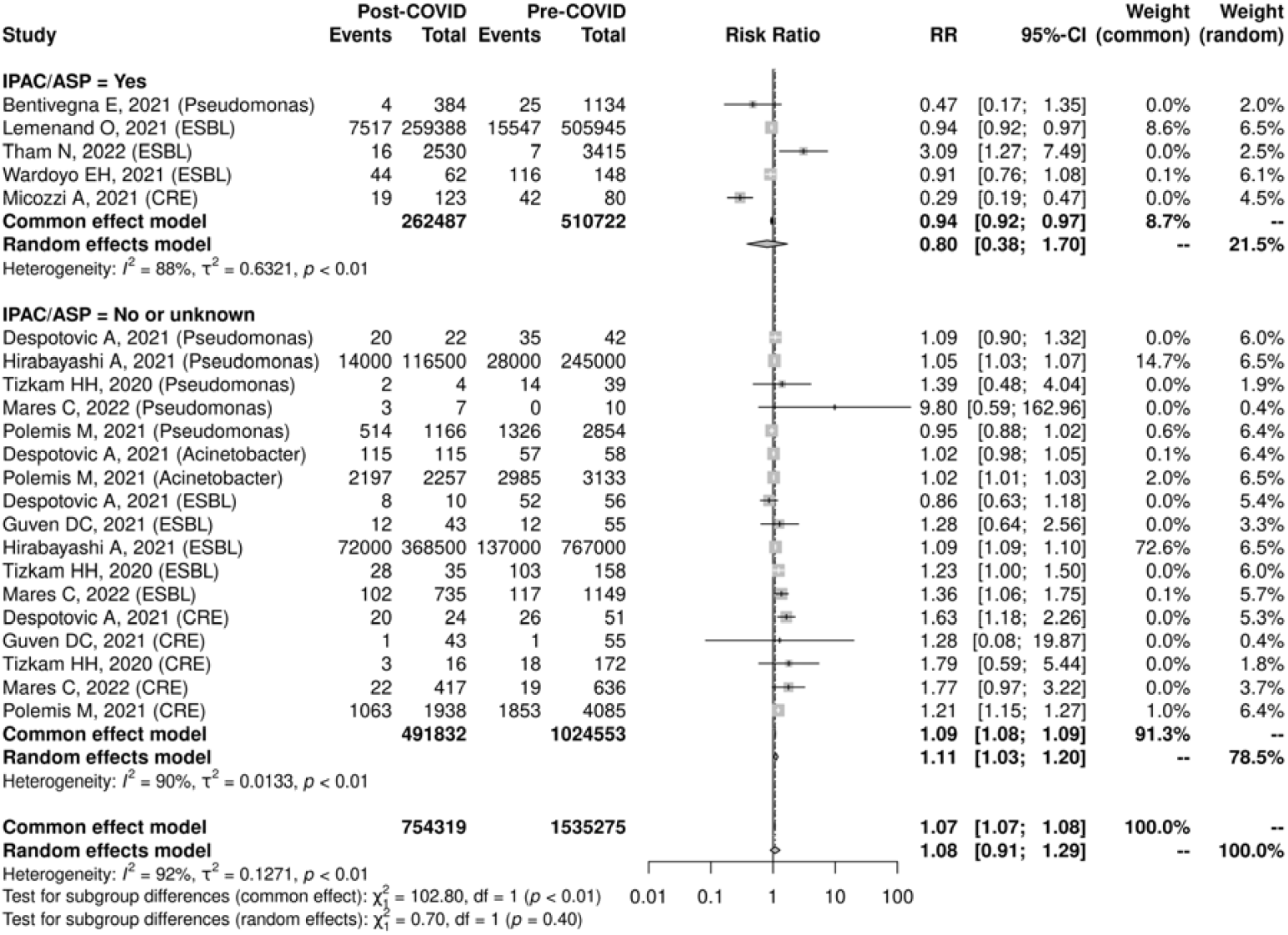
COVID-19 Pandemic and Gram-Negative AMR Risk Ratio and Presence/Absence of IPAC/ASP.

## Discussion

Antimicrobial resistance measurement in the context of the COVID-19 pandemic is variable with substantial heterogeneity in reported outcomes. We found that although the COVID-19 pandemic was not associated with a change in Gram-positive AMR, Gram-negative resistance appears to have increased (MDR or carbapenem-resistant *Pseudomonas* or *Acinetobacter*, ESBL and CRE), particularly in settings where enhanced IPAC and ASP initiatives were not reported.

A recent special report from the US Centers for Disease Control and Prevention (CDC) found a 15% increase in the rate (per discharge or admission) of resistant organisms including carbapenem-resistant *Acinetobacter*, MRSA, CRE, and ESBL.^8^ Study location, burden of COVID-19, burden of non-COVID-19 respiratory infections, and antimicrobial prescribing practices may partially explain the difference between the CDC data and our findings. The apparent increase in Gram-negative AMR but not Gram-positive AMR suggests antibiotic prescribing may play an important role, given the high use of beta-lactam/beta-lactamase inhibitors and third generation cephalosporins in patients with COVID-19.^3^ Nevertheless, both the CDC report and this systematic review present a concern that the COVID-19 pandemic may play a role in increasing rates of AMR in the population.

The concern for increasing AMR in the context of COVID-19 has been previously highlighted.^40,41^ Inappropriately high usage of antibiotics in patients with COVID-19 selects for resistant organisms which can then be transmitted to the broader population.^41^ We have previously shown that in the context of COVID-19 co-infection and secondary infections, 38% of organisms and 61% of patients harbour AMR.^4^ This analysis extends the concern for drug resistance beyond COVID-19 infected patients themselves to document an ecologic impact of the pandemic on AMR.

The relationship between COVID-19 and AMR is complex, as several factors such as improved hand hygiene, personal protective equipment use, and physical distancing may help to reduce the transmission of AMR organisms, at least temporarily while such enhanced measures are in place.^7^ Our findings reinforce that infection prevention and control activities are important mitigating factors limiting the growth of AMR associated with COVID-19.

While this study provides a broad global view of AMR in the context of COVID-19 from an ecological perspective, there are several important limitations. There is significant heterogeneity in methodology and AMR outcome measures reported across studies. At least five different AMR metrics were provided (incidence density, incidence per admission/discharge, proportion of infections, standardized infection ratio, point prevalence), which prevents direct comparison and makes meta-analysis challenging. A lack of adjustment for confounding factors raises the possibility that changes in AMR over time may be due to changes in patient populations. And lack of longitudinal data also limits the ability to account for existing temporal trends in AMR incidence and prevalence. Many studies did not comment on other confounding factors such as the presence or intensity of their infection prevention and control or antimicrobial stewardship program, so in studies which mention these initiatives, the association with reduced AMR may represent correlation rather than causation. Several studies only reported a small number of pathogens with AMR, hence there is a risk of selective outcome reporting. It is also important to note that less than 20% of studies had low risk of bias, suggesting that higher quality studies are needed to better understand the impact of COVID-19 on AMR.

## Conclusion

The COVID-19 pandemic could play an important role in the emergence and transmission of AMR, particularly for Gram-negative organisms in hospital settings. There is considerable heterogeneity in both the AMR metrics utilized and the rate of resistance reported across studies. Our findings reinforce not only the need for strengthened infection prevention and antimicrobial stewardship, but also robust and consistent AMR surveillance as part of the pandemic response and recovery.

## Data Availability

All data produced in the present study are available upon reasonable request to the authors

