## Supplemental Material for "Antibiotic Resistance associated with the COVID-19 Pandemic: A Rapid Systematic Review"

WHO COVID 19 DATABASE
<https://search.bvsalud.org/global-literature-on-novel-coronavirus-2019-ncov/>

| **#** |  | **Results**  01/12/2021 |
| --- | --- | --- |
| **1** - **Coinfection** | Co-infect* OR coinfect* OR superinfect* OR "super infection"~3 OR "super infections"~3 OR "secondary infection"~3 OR "secondary infections"~3 OR "concomitant infection"~3 OR "concomitant infections"~3 OR "mixed infection"~3 OR "mixed infections"~3 OR co-exist* OR "cross infection"~3 OR "cross infections"~3 OR polymicrobial* OR "hospital acquired" OR "healthcare associated infection" OR "healthcare associated infections" OR ab:HAI OR ti:HAI OR nosocomial* OR "ventilator associated"~3 OR superimpose* | 6,806 |
| **2 - Bacterial** | *bacteri* OR "gram negative" OR "gram positive" OR "other pathogens"~3 OR antimicrobial* OR antibiotic* OR "microbial resistant" OR "microbial resistance" OR "drug resistant" OR "drug resistance" OR "culture positive" OR "culture negative" OR sepsis OR "blood infection"~3 OR "blood infections"~3 OR "bloodstream infection"~3 OR "bloodstream infections"~3 or pyemia OR pyaemia OR "skin disease" OR "skin diseases" OR "Lower respiratory tract infection" OR LRTI OR LRTIS OR septic* | 17,963 |
| **3 bacterial syndromes** | "bacterial pneumonia"~3 OR bacteriuria* OR bacteremia* OR bacteraemia* OR "urinary infection"~3 OR "urinary infections"~3 OR UTIs OR UTI OR pyelonephritis OR cystitis OR pyuria OR cellulitis OR "soft tissue infection" OR "soft tissue infections" | 4,185 |
| **#4** | (#1 AND #2) OR (#3)  ((Co-infect* OR coinfect* OR superinfect* OR "super infection"~3 OR "super infections"~3 OR "secondary infection"~3 OR "secondary infections"~3 OR "concomitant infection"~3 OR "concomitant infections"~3 OR "mixed infection"~3 OR "mixed infections"~3 OR co-exist* OR "cross infection"~3 OR "cross infections"~3 OR polymicrobial* OR "hospital acquired" OR "healthcare associated infection" OR "healthcare associated infections" OR ab:HAI OR ti:HAI OR nosocomial* OR "ventilator associated"~3 OR superimpose*) AND (*bacteri* OR "gram negative" OR "gram positive" OR "other pathogens"~3 OR antimicrobial* OR antibiotic* OR "microbial resistant" OR "microbial resistance" OR "drug resistant" OR "drug resistance" OR "culture positive" OR "culture negative" OR sepsis OR "blood infection"~3 OR "blood infections"~3 OR "bloodstream infection"~3 OR "bloodstream infections"~3 or pyemia OR pyaemia OR "skin disease" OR "skin diseases" OR "Lower respiratory tract infection" OR LRTI OR LRTIS OR septic*)) OR ("bacterial pneumonia"~3 OR bacteriuria* OR bacteremia* OR bacteraemia* OR "urinary infection"~3 OR "urinary infections"~3 OR UTIs OR UTI OR pyelonephritis OR cystitis OR pyuria OR cellulitis OR "soft tissue infection" OR "soft tissue infections") | 6,028 |
| **Part 1:** | #4 AND Entry_date:([20190101 TO 20210201]) | 2.816 |
| **Part 2:** | #4 AND Entry_date:([20210201 TO 20211201]) | 3220 |
